# SARS-CoV-2 Antibody Testing in Healthcare Workers: a comparison of the clinical performance of three commercially available antibody assays

**DOI:** 10.1101/2021.05.25.21257772

**Authors:** Niamh Allen, Melissa Brady, Antonio Isidro Carrion Martin, Lisa Domegan, Cathal Walsh, Elaine Houlihan, Colm Kerr, Lorraine Doherty, Joanne King, Martina Doheny, Damian Griffin, Maria Molloy, Jean Dunne, Vivion Crowley, Philip Holmes, Evan Keogh, Sean Naughton, Martina Kelly, Fiona O’Rourke, Yvonne Lynagh, Brendan Crowley, Cillian de Gascun, Paul Holder, PRECISE Study Steering Group, Colm Bergin, Catherine Fleming, Una Ni Riain, Niall Conlon

## Abstract

SARS-CoV-2 antibodies are an excellent indicator of past COVID-19 infection. As the COVID-19 pandemic progresses, retained sensitivity over time is an important quality in an antibody assay that is to be used for the purpose of population seroprevalence studies.

We compared 5788 healthcare worker (HCW) serum samples on two serological assays (Abbott SARS-CoV-2 anti-nucleocapsid IgG and Roche Anti-SARS-CoV-2 anti-nucleocapsid Total Antibody) and a subset of samples (all Abbott assay positive or grayzone, n=485) on Wantai SARS-CoV-2 anti-spike Antibody ELISA. For 367 samples from HCW with previous PCR-confirmed SARS-CoV-2 infection we correlated the timing of infection with assay results.

Overall seroprevalence was 4.2% on Abbott, 9.5% on Roche. Of those with previously confirmed infection, 41% (150/367) and 95% (348/367) tested positive on Abbott and Roche respectively. At 21 weeks (150 days) after confirmed infection, positivity on Abbott started to decline. Roche positivity was retained for the entire study period (33 weeks). Factors associated (*P*≤ 0.050) with Abbott seronegativity in those with previous PCR-confirmed infection included sex (male OR0.30;95%CI0.15-0.60), symptom severity (OR0.19 severe symptoms;95%CI0.05-0.61), ethnicity (OR0.28 Asian ethnicity;95%CI0.12-0.60) and time since PCR diagnosis (OR2.06 for infection 6 months previously;95%CI1.01-4.30. Wantai detected all previously confirmed infections.

In our population, Roche detected antibodies up to at least seven months after natural infection with SARS-CoV-2. This may indicate that Roche is better suited than Abbott to population-based studies. Wantai demonstrated high sensitivity but sample selection was biased. The relationship between serological response and functional immunity to SARS-CoV-2 infection needs to be delineated.

## Introduction

Serological assays for the detection of severe acute respiratory syndrome coronavirus-2 (SARS-CoV-2) antibodies have a significant role to play in the response to the COVID-19 pandemic worldwide. Detectable antibody to SARS-CoV-2 is an excellent indicator of past SARS-CoV-2 infection (1) and therefore helps determine the proportion of a population that has been previously exposed or infected. Population seroprevalence studies can provide crucial supplementary information to national surveillance systems and have the benefit of accounting for asymptomatic cases and symptomatic individuals who may not present to the health services for testing. As mass COVID-19 vaccination programmes are rolled out globally, serological assays may have a role in assessing host vaccine response and predicting vaccine effectiveness (2). They may also potentially be used to inform individual risk of disease (3) with emerging evidence of up to six months of immunity after natural infection (4), though further research on this area of post-infection immunity is required.

Many studies have examined the timing of antibody production and sensitivity of commercially available antibody assays in the early stages of infection with SARS-CoV-2 (5) (6). Few studies have evaluated and compared the longevity of assay sensitivity post PCR-confirmed infection for different assays on the same HCW population. As the pandemic progresses, retained sensitivity over time is an important quality in an antibody assay that is to be used for the purpose of population seroprevalence studies. Decline in SARS-CoV-2 antibody response to natural infection over time has been described (7) (8), and that different antibodies may wane at different rates. while other studies have shown sustained antibody detection for up to 100-125 days (9) (10). Understanding the duration of antibody response over time is key to understanding the accuracy of epidemiological studies and informing public health pandemic response measures. The extent and duration of immunity and its relationship to antibody positivity is not yet fully understood, however the use of serology at individual level for the purpose of estimating immunity also requires detailed understanding of the timing and waning of antibody response in relation to PCR-positivity. A UK study comparing five widely available commercial assays (including the Abbott Architect IgG assay and Roche Elecsys Anti-SARS-CoV-2 assay) found them all to have a sensitivity and specificity of at least 98% 30 days post symptom onset (11). Regarding longevity of the antibody response there are few published data, and results differ; a study comparing four different serological assays showed a decline in the performance of the Abbott assay after 60 days, and a further decline after 80 days (12), whereas another study of the same assay showed a mean of 137 days to loss of positive antibody (13). Further data comparing antibody assays for the detection of antibodies to SARS-CoV-2 are needed to guide the most appropriate use of certain commercially available assays in any given situation.

The primary purpose of the data collected was to estimate the seroprevalence of past SARS-CoV-2 infection in our HCW population; that analysis was carried separately as part of the PRECISE Study on Prevalence of Antibodies to SARS-CoV-2 in Irish Healthcare Workers (14). The data presented here is a secondary analysis as part of the PRECISE study, with the aim of comparing the prevalence of anti-SARS-CoV-2 antibodies in the same HCW population in two different assays targeting the anti-nucleocapsid protein; Abbott Architect SARS-CoV-2 immunoglobulin (Ig)G assay and Roche Elecsys Anti-SARS-CoV-2 immunoassay (15) (16) (17). We aimed to also compare antibody prevalence for a subset of samples on an assay targeting the anti-spike protein; the Wantai SARS-CoV-2 Antibody ELISA.

## Methods

### Study Design

This was a cross-sectional study of the seroprevalence of circulating antibodies to SARS-CoV-2 in hospital HCW, performed in October 2020 (18). All staff members of both hospitals were invited to participate in an online self-administered consent process and online questionnaire, followed by blood sampling for SARS-CoV-2 antibody testing. Information collected in the questionnaire included demographic information and information regarding previous COVID-19 symptoms, testing and diagnosis.

### Laboratory Assays

All samples were tested by two assays; the Abbott SARS-CoV-2 measuring IgG (referred to here as Abbott) and Roche Elecsys Anti-SARS-CoV-2 measuring total antibodies (referred to here as Roche) (15) (16) (17). The Abbott SARS-CoV-2 IgG assay (Abbott) is a chemiluminescent microparticle immunoassay that detects IgG antibodies to the nucleocapsid protein of SARS-CoV-2. The Roche Elecsys Anti-SARS-CoV-2 total antibody assay (Roche) is an electrochemiluminescent immunoassay that detects antibodies (including IgG), also to the nucleocapsid protein.

Assay results were interpreted using the manufacturers’ recommended assay specific thresholds. Assay threshold ≥ 1.4 (sample to calibrator (S/C) index) for Abbott and ≥ 1.0 (Cutoff index (COI)) for Roche were determined to be reactive and interpreted as antibody positive. The Abbott SARS-CoV-2 IgG grayzone is an additional assay threshold band for potential positivity, suggested by the manufacturer to increase assay sensitivity (Abbott Diagnostics Product Information Letter PI1060-2020) (16), see Table 1 for interpretation of the Abbott S/C index within this study. All samples with an Abbott result of positive or grayzone were tested on a third assay in the National Virus Reference Laboratory (NVRL) using the Wantai SARS-CoV-2 Antibody ELISA (referred to here as Wantai), distributed by Fortress Diagnostics. Wantai is an Enzyme-Linked Immunosorbent Assay (ELISA) for qualitative detection of total antibodies (including IgG and IgM) to the spike protein of SARS-CoV-2.

**Table 1.**
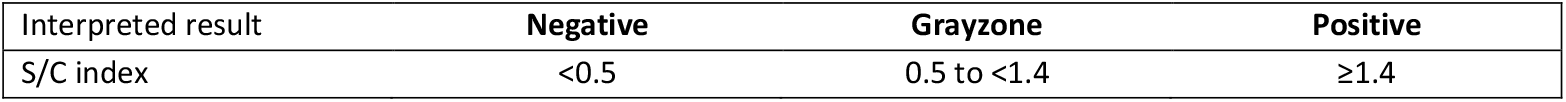
Interpretation of the Abbott S/C index

| Interpreted result | Negative | Grayzone | Positive |
| --- | --- | --- | --- |
| S/C index | <0.5 | 0.5 to <1.4 | ≥1.4 |

In terms of assay performance, according to the manufacturer’s specifications all three assays perform with high sensitivity and high specificity, see Table 2. (19).

**Table 2.** Summary of assay performance according to manufacturer specifications Sensitivity

|  | Sensitivity | Specificity |
| --- | --- | --- |
| **Abbott SARS-CoV-2 IgG** | 100% (95% CI; 95.8 - 100) | 99.6% (95% CI 99.0 - 99.9) |
| **Roche Anti-SARS-CoV-2** | 100% (95% CI; 88.3 - 100) | 99.8% (95% CI 99.7 - 99.9) |
| **Wantai SARS-CoV-2 Antibody** | 96.7% (95% CI 83.3 - 99.4) | 97.5% (95% CI 91.3 - 99.3) |

### Statistical methods

Assay concordance was assessed using Cohen’s kappa statistic and McNemar’s test for difference in proportions. Cohen’s kappa coefficient (κ) measures the level of agreement between assays, taking into account the possibility of the agreement occurring by chance. The κ statistic varies from 0 to 1, where 0 = agreement equivalent to chance, 0.1 – 0.20 = slight agreement, 0.21 – 0.40 = fair agreement, 0.41 – 0.60 = moderate agreement, 0.61 – 0.80 = substantial agreement, 0.81 – 0.99 = near perfect agreement and 1 = perfect agreement.

Confidence intervals (CI) for the proportion of participants that were seropositive were computed. Multivariable logistic regression was carried out to assess risk factors for the absence of SARS-CoV-2 IgG antibodies controlling for age, sex, ethnicity/background, type of patient contact, severity of symptoms and number of months since PCR positive test. Forward stepwise selection was used, the Akaike information criterion (AIC) was used to evaluate model efficiency; HCW role was excluded in the final model. We used R version 4.0.3 (R Foundation for Statistical Computing, Vienna, Austria) and OpenEpi software version 3.01 (Wilson Score) (20).

## Results

COVID-19 serology test results were available for 5,788 participating HCWs (comprising 64% of all staff in two Irish tertiary referral hospitals). The majority of participants were female (77%); median age was 39 years (IQR 30-49). A small proportion (5%) of participants were over 60 years of age. By role, the highest proportion of participants were nursing staff (36%). Characteristics of participants by serology assay are shown in Appendix 1A and 1B.

### SARS-CoV-2 seropositivity in relation to assay type: Abbott SARS-CoV-2 and Roche Anti-SARS-CoV-2

All 5,788 participants were tested using the Abbott SARS-CoV-2 IgG assay and 99.9% (n=5,787) were tested using the Roche Anti-SARS-CoV-2 assay. A considerably lower proportion of participants had a positive antibody on the Abbott assay (4.2%) compared to the proportion that had a positive antibody on the Roche assay (9.5%) (Table 3).

**Table 3.**
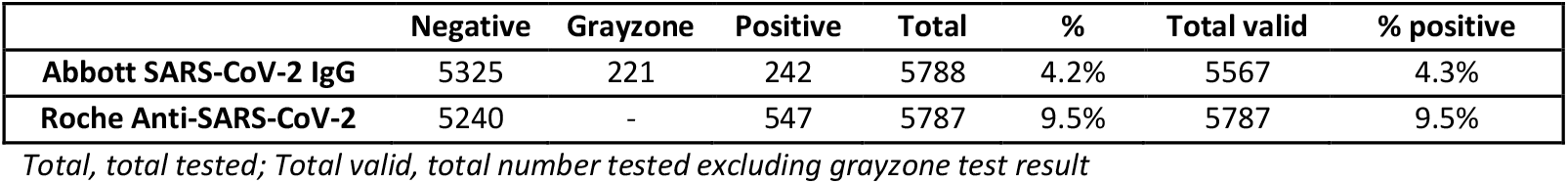
Serology result by assay

Assay concordance was moderate (k -0.53; 95% CI: 0.48 to 0.57) (Table 4). McNemar’s test for difference in proportions indicated a systematic difference in the proportion of positive results between the two assays (p<0.001). Twenty-four participants tested positive on Abbott but negative on Roche. Of the 24, three also tested positive on the Wantai assay, suggesting possible false negative results on Roche for these three participants. Of these three, one was recently diagnosed with COVID-19 (positive by PCR 16 days prior to serology testing). Of the remaining 21, all tested negative on Wantai; none had had prior PCR-confirmed SARS-CoV-2 infection; 11/21 had never been tested and 10/21 had had a negative COVID-19 PCR test at some stage. Of the 21, nine (43%) reported ever having symptoms of COVID-19 (indicating possible undiagnosed infection), eight of whom had mild symptoms (similar to a cold or less) and one of whom had significant symptoms (similar to influenza but not requiring hospital admission). The interval between date of previous symptoms and date of serology testing varied; less than one month (n=1), one month (n=1), two months (n=2), three months (n=1), five months (n=1), six months (n=1), seven months (n=2). Analysis was carried out in order to explore the association between participant characteristics and discordant results between the Abbott and Roche assays, there was no significant association observed (data not presented here).

**Table 4.**
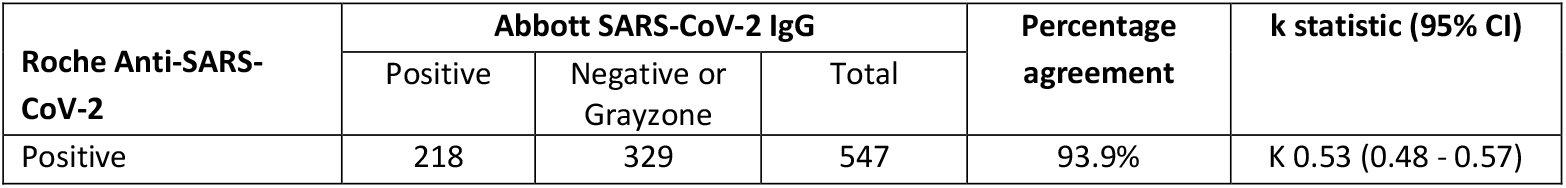
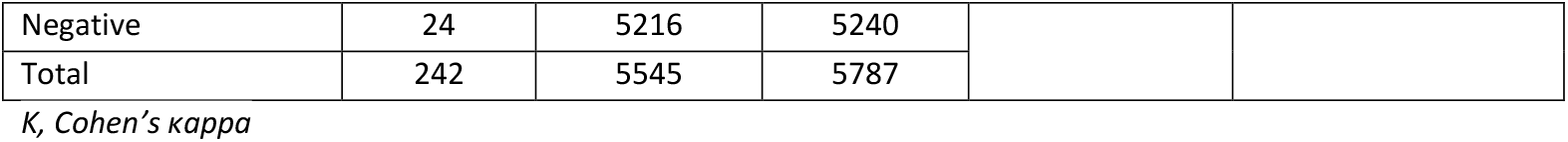
Distribution of positive and negative results by serology assay

| Roche Anti-SARS-CoV-2 | Abbott SARS-CoV-2 IgG |  |  | Percentage agreement | k statistic (95% CI) |
| --- | --- | --- | --- | --- | --- |
|  | Positive | Negative or Grayzone | Total |  |  |
| Positive | 218 | 329 | 547 | 93.9% | K 0.53 (0.48 - 0.57) |
| Negative | 24 | 5216 | 5240 |  |  |
| Total | 242 | 5545 | 5787 |  |  |

*K, Cohen's kappa*

### Abbott SARS-CoV-2 ‘Grayzone’ results

Four percent of participants (n=221) had results in the Abbott SARS-CoV-2 IgG grayzone. Of those 67% (n=149) tested positive on the Roche assay. Of those with grayzone results, 42% were previously diagnosed with COVID-19 (positive by PCR). There were no participant characteristics that were significantly associated with having grayzone test results.

In order to explore whether the numerical value within the grayzone S/C index range could be used to assist interpretation of the grayzone, arbitrary cut-offs (low, medium, high) were applied, and these were compared to the interpreted results on the Roche assay. There was no correlation observed, as a similar proportion of results within each of the arbitrary cut-offs were positive on the Roche assay (Table 5).

**Table 5.**
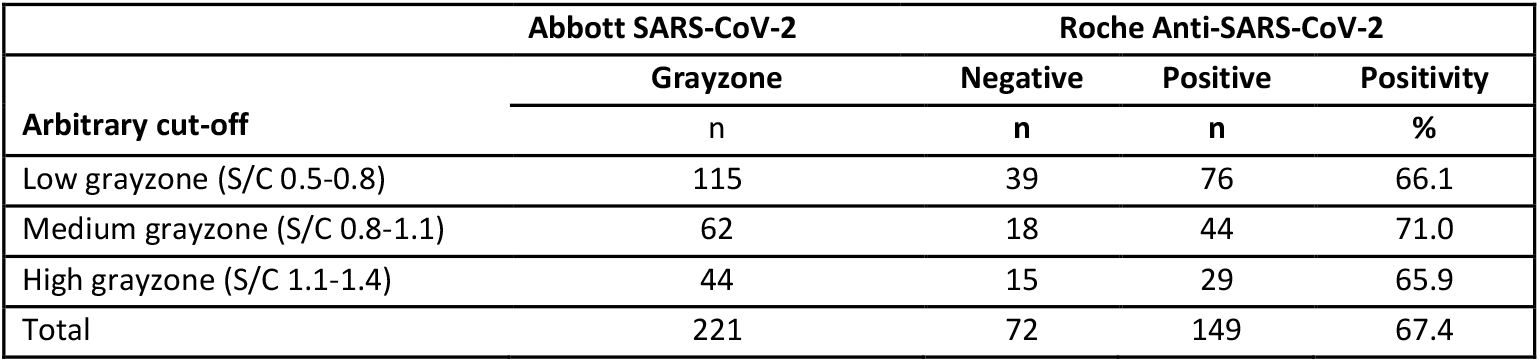
Comparison of Abbott SARS-CoV-2 IgG S/C index and Roche Anti-SARS-CoV2 interpreted result, among participants with grayzone results (n=221)

### SARS-CoV-2 seropositivity in relation to assay type: Roche Anti-SARS-CoV-2 and Wantai SARS-CoV-2 Ab ELISA

In total, 8.4% (n=485) of participants were tested using both the Roche assay and the Wantai assay. Assay concordance was almost perfect (k -0.93; 95% CI: 0.88 to 0.97) (Table 6). There was no evidence of difference in the proportion of positive results between the two assays (p<0.131).

**Table 6.**
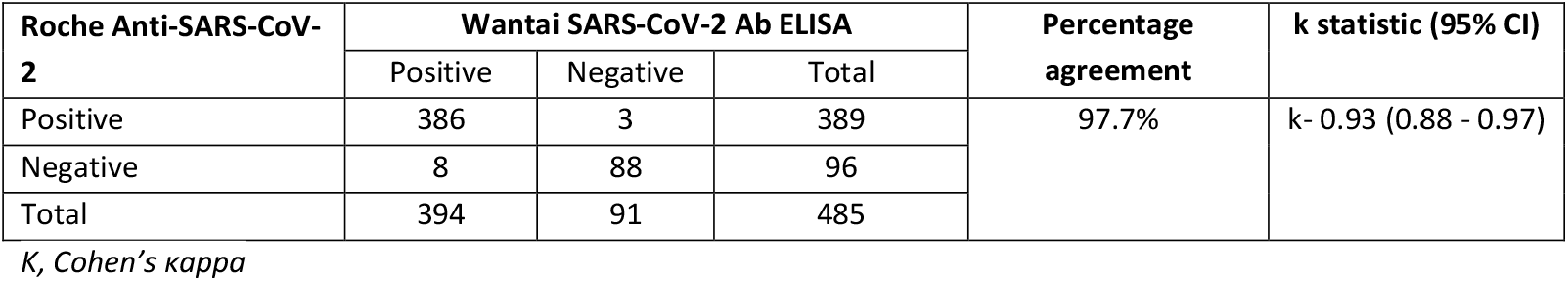
Distribution of positive and negative results by assay

### SARS-CoV-2 seropositivity over time (previously PCR-positive participants)

Three hundred and sixty-seven participants were previously diagnosed COVID-19 positive by PCR. Among those, 41% (150/367) (95% CI 35-46) tested positive on Abbott and 95% (348/367) (95% CI 92-96) tested positive on Roche. There were 93 participants who were previously PCR-positive who had a result in the Abbott grayzone. If the Abbott grayzone was included in positive results, that would increase Abbott positivity to 66% (243/367).

Two-hundred and fifty-nine (71%) previously PCR-positive participants were tested on Wantai, and all 259 had a positive result on the Wantai assay. Results for the Wantai assay are excluded from this section as not all samples were tested on this platform.

The self-reported date of previous positive PCR test was available for 365 (99%) participants. The interval between date of previous positive PCR test and date of serology test ranged from 12 to 231 days (2-33 weeks; 0-7 months). Serology test result by number of weeks since positive PCR test (including the breakdown for those who were symptomatic at the time of PCR testing) is shown in Table 7 and visually represented in Figure 1a, b and c (visual representation excludes participants with grayzone results). We saw a decline in antibody positivity on the Abbott assay from week 21 (day 150) onwards. Figure 2 shows the percentage positivity by time (months) since PCR test, including 95% confidence intervals (CI), showing a decline in antibody positivity on the Abbott assay in month 4.

**Table 7.** Detection of SARS-CoV-2 antibodies by serology assay type with respect to time (number of weeks) since positive PCR test

| N<br>weeks | Previously PCR positive (n=367) |  |  |  |  |  |  |  |  |  | Previously PCR positive, and had symptoms at time of PCR test (n=340) |  |  |  |  |  |  |  |  |  |
| --- | --- | --- | --- | --- | --- | --- | --- | --- | --- | --- | --- | --- | --- | --- | --- | --- | --- | --- | --- | --- |
|  | Abbott SARS-CoV-2 IgG |  |  |  |  |  | Roche Anti-SARS-CoV-2 |  |  |  | Abbott SARS-CoV-2 IgG |  |  |  |  |  | Roche Anti-SARS-CoV-2 |  |  |  |
|  | N | G | P | T | %<br>positive | %<br>positive<br>(valid<br>test) | N | P | T | %<br>positive | N | G | P | T | %<br>positive | %<br>positive<br>(valid<br>test) | N | P | T | %<br>positive |
| 2 | 0 | 0 | 5 | 5 | 100 | 100 | 1 | 4 | 5 | 80.0 | 0 | 0 | 2 | 2 | 100 | 100 | 1 | 1 | 2 | 50.0 |
| 3 | 1 | 0 | 0 | 1 | 0.0 | 0.0 | 1 | 0 | 1 | 0.0 | 1 | 0 | 0 | 1 | 0.0 | 0.0 | 1 | 0 | 1 | 0.0 |
| 4 | 0 | 1 | 2 | 3 | 66.7 | 100 | 1 | 2 | 3 | 66.7 | 0 | 1 | 2 | 3 | 66.7 | 100 | 1 | 2 | 3 | 66.7 |
| 5 | 0 | 0 | 4 | 4 | 100 | 100 | 0 | 4 | 4 | 100 | 0 | 0 | 4 | 4 | 100 | 100 | 0 | 4 | 4 | 100 |
| 6 | 0 | 0 | 1 | 1 | 100 | 100 | 0 | 1 | 1 | 100 | 0 | 0 | 1 | 1 | 100 | 100 | 0 | 1 | 1 | 100 |
| 7 | 0 | 0 | 2 | 2 | 100 | 100 | 0 | 2 | 2 | 100 | 0 | 0 | 2 | 2 | 100 | 100 | 0 | 2 | 2 | 100 |
| 9 | 0 | 0 | 1 | 1 | 100 | 100 | 0 | 1 | 1 | 100 | 0 | 0 | 1 | 1 | 100 | 100 | 0 | 1 | 1 | 100 |
| 10 | 0 | 0 | 2 | 2 | 100 | 100 | 0 | 2 | 2 | 100 | 0 | 0 | 2 | 2 | 100 | 100 | 0 | 2 | 2 | 100 |
| 11 | 0 | 0 | 1 | 1 | 100 | 100 | 0 | 1 | 1 | 100 | 0 | 0 | 0 | 0 | 100 | 100 | 0 | 0 | 0 | 100 |
| 17 | 0 | 0 | 1 | 1 | 100 | 100 | 0 | 1 | 1 | 100 | 0 | 0 | 1 | 1 | 100 | 100 | 0 | 1 | 1 | 100 |
| 19 | 0 | 0 | 2 | 2 | 100 | 100 | 0 | 2 | 2 | 100 | 0 | 0 | 2 | 2 | 100 | 100 | 0 | 2 | 2 | 100 |
| 20 | 0 | 0 | 1 | 1 | 100 | 100 | 0 | 1 | 1 | 100 | 0 | 0 | 1 | 1 | 100 | 100 | 0 | 1 | 1 | 100 |
| 21 | 0 | 1 | 5 | 6 | 83.3 | 100 | 0 | 6 | 6 | 100 | 0 | 1 | 5 | 6 | 83.3 | 100 | 0 | 6 | 6 | 100 |
| 22 | 0 | 4 | 7 | 11 | 63.6 | 100 | 0 | 11 | 11 | 100 | 0 | 3 | 6 | 9 | 66.7 | 100 | 0 | 9 | 9 | 100 |
| 23 | 3 | 1 | 7 | 11 | 63.6 | 70.0 | 1 | 10 | 11 | 90.9 | 2 | 1 | 7 | 10 | 70.0 | 77.8 | 0 | 10 | 10 | 100 |
| 24 | 6 | 5 | 6 | 17 | 35.3 | 50.0 | 0 | 17 | 17 | 100 | 5 | 5 | 5 | 15 | 33.3 | 50.0 | 0 | 15 | 15 | 100 |
| 25 | 9 | 12 | 12 | 33 | 36.4 | 57.1 | 0 | 33 | 33 | 100 | 9 | 11 | 12 | 32 | 37.5 | 57.1 | 0 | 32 | 32 | 100 |
| 26 | 7 | 10 | 7 | 24 | 29.2 | 50.0 | 1 | 23 | 24 | 95.8 | 7 | 9 | 7 | 23 | 30.4 | 50.0 | 1 | 22 | 23 | 95.7 |
| 27 | 18 | 7 | 23 | 48 | 47.9 | 56.1 | 2 | 46 | 48 | 95.8 | 16 | 7 | 21 | 44 | 47.7 | 56.8 | 2 | 42 | 44 | 95.5 |
| 28 | 20 | 9 | 18 | 47 | 38.3 | 47.4 | 2 | 45 | 47 | 95.7 | 19 | 8 | 17 | 44 | 38.6 | 47.2 | 1 | 43 | 44 | 97.7 |
| 29 | 20 | 17 | 24 | 61 | 39.3 | 54.5 | 6 | 55 | 61 | 90.2 | 19 | 16 | 24 | 59 | 40.7 | 55.8 | 6 | 53 | 59 | 89.8 |
| 30 | 31 | 18 | 9 | 58 | 15.5 | 22.5 | 4 | 54 | 58 | 93.1 | 29 | 18 | 8 | 55 | 14.5 | 21.6 | 4 | 51 | 55 | 92.7 |
| 31 | 8 | 7 | 6 | 21 | 28.6 | 42.9 | 0 | 21 | 21 | 100 | 6 | 7 | 6 | 19 | 31.6 | 50.0 | 0 | 19 | 19 | 100 |
| 32 | 0 | 1 | 2 | 3 | 66.7 | 100 | 0 | 3 | 3 | 100 | 0 | 1 | 2 | 3 | 66.7 | 100 | 0 | 3 | 3 | 100 |
| 33 | 1 | 0 | 0 | 1 | 0.0 | 0.0 | 0 | 1 | 1 | 100 | 1 | 0 | 0 | 1 | 0.0 | 0.0 | 0 | 1 | 1 | 100 |
| Total | 124 | 93 | 150 | 367 | 40.9 | 54.7 | 19 | 348 | 367 | 94.8 | 114 | 88 | 138 | 340 | 40.6 | 54.8 | 17 | 323 | 340 | 95.0 |

Serology test results = N, number negative; G, number grayzone; P, number positive; T, total tested; % positive valid test excluded grayzone results. Table 7
excludes two participants for which time since positive PCR test was unknown.

**Figure 1.**
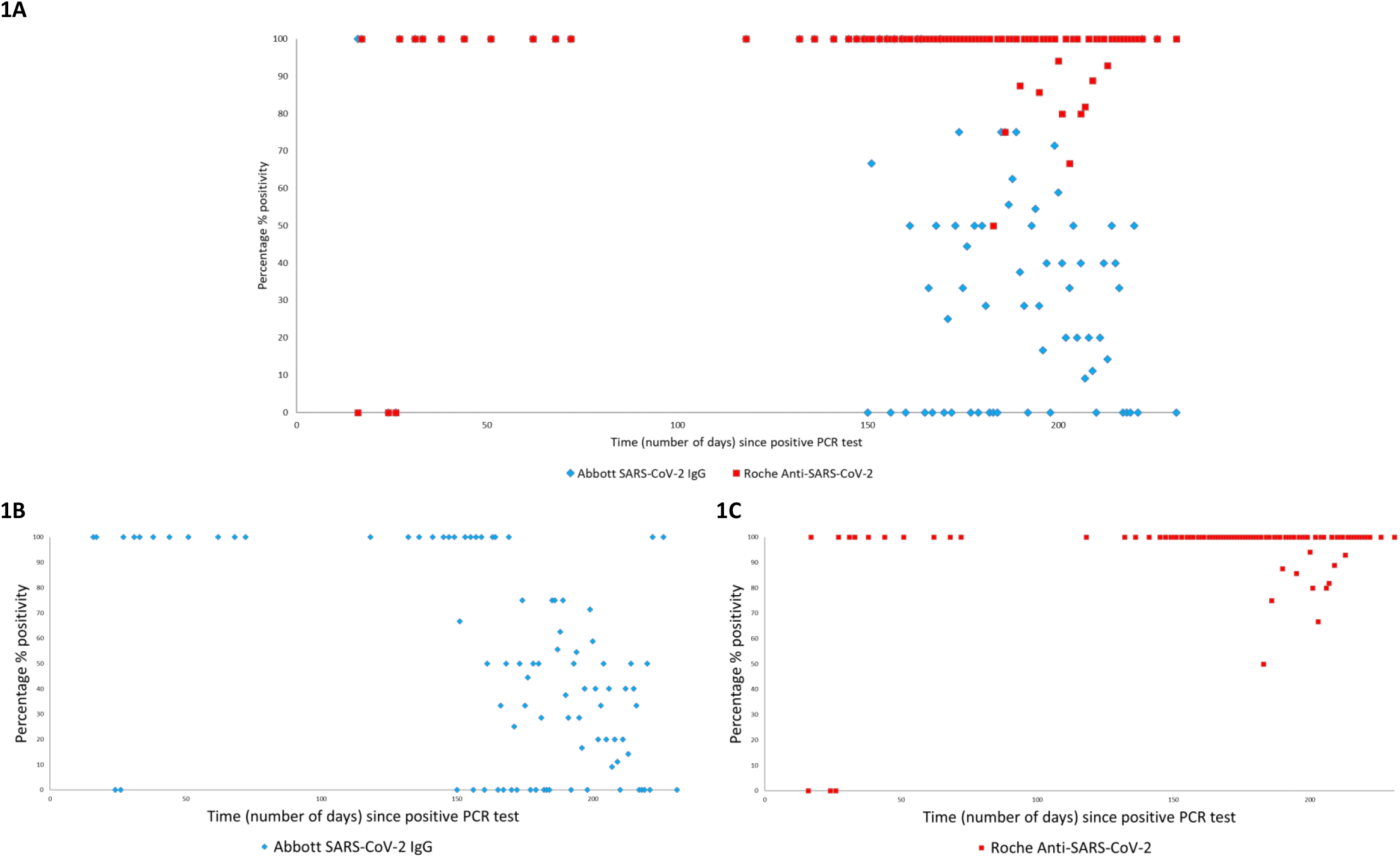
**A-C** Percentage of participants with detected SARS-CoV-2 antibodies by serology assay with respect to time (number of days) since positive PCR test, among participants who had previous PCR-confirmed COVID-19 infection and had symptoms at the time of their PCR test (n=340). Figure **1A** both assays, **1B** Abbott only, **1C** Roche only.

**Figure 2.**
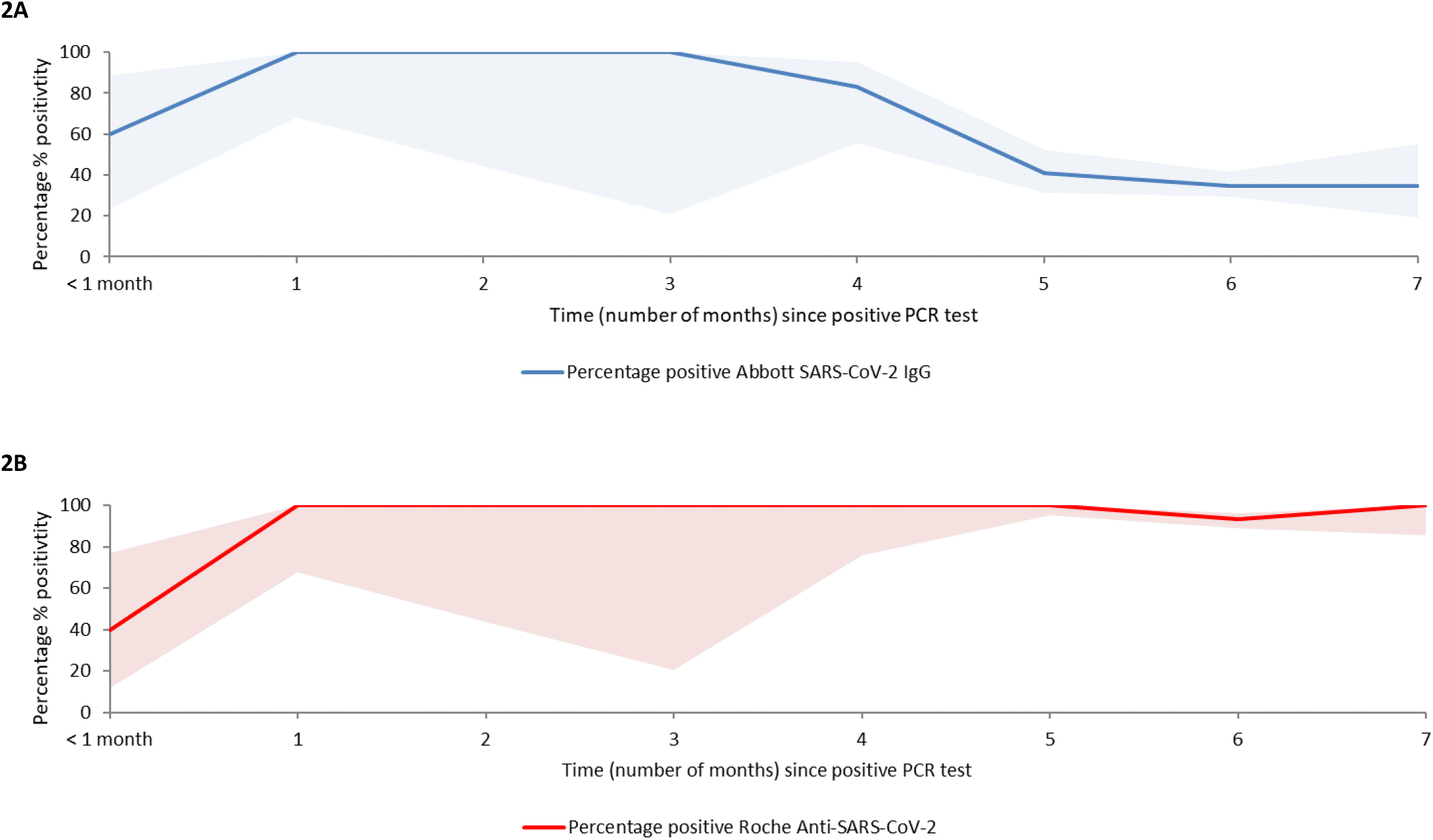
**A and 2B** Percentage % positivity among participants by serology assay with respect to time (number of months) since positive PCR test, among participants who had previous PCR-confirmed SARS-CoV-2 infection and had symptoms at the time of their PCR test (n=340). Shaded area indicates 95% confidence interval (CIs). Note: CIs are wide for months 0-4 and month 7 due to a low number of participants at these testing intervals. Figure **2A** (Abbott only), **2B** (Roche only)

The most common interval between positive PCR test and serology testing was six months (61%; n=222). There were 210 participants who had a six-month PCR to serology testing interval, and who were symptomatic at the time of their PCR test; among them positivity was 35% (95% CI 29-41) on Abbott (n=73) and it was 93% (95% CI 89-96) for Roche (n=196).

Seventeen participants (4.6%) with a previously PCR-confirmed infection had negative serology test on both the Abbott and Roche assays, and also tested negative on the Wantai assay. Of the 17, one had recent COVID-19 infection (24 days prior) and the remaining 16 had distant infection (positive PCR was five (n=1) or six (n=15) months prior to serology testing). Characteristics of the 17 participants are shown in Appendix 2. Among the 17, a lower proportion (88%; n=15) had symptoms at the time of their PCR positive test when compared to the overall PCR-positive subgroup (98%; n=358). A higher proportion were of white Irish background (94% versus 65% in the overall PCR-positive subgroup). Other characteristics did not differ considerably. Analysis was carried out in order to explore the association between participant characteristics and negative serology results; there was no significant association observed (data not presented here).

In order to explore whether the Roche quantitative COI or the Abbott S/C index was close to the positive threshold for these 17 participants, arbitrary cut-offs (low, medium and high negative) were applied to the results of both assays. For the Roche assay, three participants had results that were close to the positive result threshold (i.e. in the high negative range: COI 0.6-0.9), six had results in the medium negative range (COI 0.3-0.6) and eight had results in the low negative range (COI 0-0.3). For the Abbott assay, all participants had results in either the medium negative range (n=3) or the low negative range (n= 14).

### Abbott SARS-CoV-2 seronegativity in relation to participant characteristics

The characteristics of participants with previous PCR-confirmed SARS-CoV-2 infection (n=367) and their serology test results by Abbott SARS-CoV-2 are shown in Appendix 3A and 3B. In total, 45% (n=124) (34% including those who had grayzone results in the total number tested) of participants with previous PCR-confirmed SARS-CoV-2 infection did not have detectable antibodies on the Abbott assay. Univariable and multivariable analysis were carried to out to explore the association between participant characteristics and negative Abbott test result. Table 8 present the results of the analysis. Factors associated with Abbott seronegativity in those with a previous PCR-confirmed infection varied by sex (male adjusted OR (aOR) 0.30; 95% CI 0.15-0.60, *P* <0.001), symptom severity (severe symptoms requiring hospitalisation: aOR 0.19; 95% CI 0.05-0.61, *P* 0.008), ethnicity (Asian ethnicity: aOR 0.28; 95% CI 0.12-0.60, *P* 0.001) and time since PCR diagnosis (increase from five-to six-month interval: aOR 2.06; 95% CI 1.01-4.30, *P* 0.050).

**Table 8.** Factors associated with Abbott SARS-CoV-2 seronegativity, among participants with previous PCR-confirmed SARS-CoV-2 infection (n=274 participants with valid results)

|  |  | All<br>(n) | Negative<br>(n) | Negative<br>% | Unadjusted<br>OR | 95% CI | P-value | Adjusted<br>OR | 95% CI | P-value |
| --- | --- | --- | --- | --- | --- | --- | --- | --- | --- | --- |
| **Total** |  | 274 | 124 | 45.3 |  |  |  |  |  |  |
| **Age group (in years)** | 18-29 | 79 | 41 | 51.9 | * |  |  | * |  |  |
|  | 30-39 | 79 | 43 | 54.4 | 1.11 | 0.59 - 2.07 | 0.750 | 0.99 | 0.46 - 2.15 | 0.982 |
|  | 40-49 | 59 | 17 | 28.8 | **0.38** | **0.18 - 0.76** | **0.007** | **0.36** | **0.15 - 0.82** | **0.017** |
|  | 50-59 | 45 | 19 | 42.2 | 0.68 | 0.32 - 1.41 | 0.301 | 0.70 | 0.28 - 1.71 | 0.429 |
|  | 60+ | 12 | 4 | 33.3 | 0.46 | 0.12 - 1.60 | 0.238 | 0.25 | 0.05 - 1.04 | 0.065 |
| **Sex** | Female | 201 | 102 | 50.7 | * |  |  | * |  |  |
|  | Male | 73 | 22 | 30.1 | **0.42** | **0.25 - 0.74** | **0.003** | **0.30** | **0.15 - 0.60** | **<0.001** |
| **Ethnicity** | Irish | 173 | 94 | 54.3 | * |  |  | * |  |  |
|  | Any other white background | 35 | 12 | 34.3 | 0.44 | 0.20 - 0.92 | 0.033 | 0.49 | 0.2 - 1.19 | 0.118 |
|  | Any Asian background | 56 | 14 | 25.0 | **0.28** | **0.14 - 0.54** | **<0.001** | **0.28** | **0.12 - 0.60** | **0.001** |
|  | Any African/black background | 7 | 2 | 28.6 | 0.34 | 0.04 - 1.61 | 0.200 | 0.47 | 0.05 - 3.33 | 0.449 |
|  | Other | 3 | 2 | 66.7 | 1.68 | 0.15 - 36.6 | 0.067 | 2.33 | 0.14 - 70.6 | 0.566 |
| **Close contact with a case of COVID-19\*\*** | Yes | 155 | 65 | 41.9 | 0.71 | 0.44 - 1.15 | 0.178 | 0.63 | 0.34 - 1.16 | 0.137 |
|  | No | 117 | 59 | 50.4 | * |  |  | * |  |  |
| **Main type of patient contact^** | No patient contact | 46 | 24 | 52.2 | * |  |  | * |  |  |
|  | Daily contact non-COVID-19 | 162 | 74 | 45.7 | 0.77 | 0.39 - 1.49 | 0.437 | 0.79 | 0.34 - 1.80 | 0.577 |
|  | Daily contact known or | 66 | 26 | 39.4 | 0.60 | 0.27 - 1.27 | 0.182 | 0.83 | 0.32 - 2.14 | 0.694 |
| **Severity of symptoms\*\*** | No symptoms | 6 | 3 | 50.0 | 0.82 | 0.15 - 4.61 | 0.816 | 0.86 | 0.12 - 6.29 | 0.876 |
|  | Minor symptoms | 113 | 62 | 54.9 | * |  |  | * |  |  |
|  | Significant symptoms | 130 | 55 | 42.3 | **0.60** | **0.36 - 1.00** | **0.051** | **0.50** | **0.27 - 0.92** | **0.028** |
|  | Severe symptoms (hospitalised) | 23 | 4 | 17.4 | **0.17** | **0.04 - 0.49** | **0.003** | **0.19** | **0.05 - 0.61** | **0.008** |
| **HCW Role** | Administration | 18 | 9 | 50.0 | * |  |  |  |  | not entered |
|  | Allied health care | 34 | 19 | 55.9 | 1.27 | 0.40 - 4.03 | 0.686 |  |  |  |
|  | General Support | 10 | 5 | 50.0 | 1.00 | 0.21 - 4.81 | 1.00 |  |  |  |
|  | Healthcare assistant | 24 | 5 | 20.8 | 0.26 | 0.06 - 0.98 | 0.053 |  |  |  |
|  | Medical/Dental | 62 | 29 | 46.8 | 0.88 | 0.30 - 2.54 | 0.809 |  |  |  |
|  | Nursing/Midwifery | 124 | 55 | 44.4 | 0.80 | 0.29 - 2.17 | 0.653 |  |  |  |
|  | Other | 2 | 2 | 100.0 | NA | NA | NA |  |  |  |
| **Number of months** | Less than one month | 7 | 1 | 14.3 | 0.25 | 0.01 - 1.57 | 0.21 | 0.11 | 0.01 - 0.83 | 0.061 |
| since PCR positive test** | One month | 8 | 0 | 0.0 | NA | NA | NA | NA | NA | NA |
|  | Two months | 4 | 0 | 0.0 | NA | NA | NA | NA | NA | NA |
|  | Three months | 1 | 0 | 0.0 | NA | NA | NA | NA | NA | NA |
|  | Four months | 10 | 0 | 0.0 | NA | NA | NA | NA | NA | NA |
|  | Five months | 57 | 23 | 40.4 | * |  |  | * |  |  |
|  | Six months | 168 | 91 | 54.2 | **1.75** | **0.95 - 3.25** | **0.073** | **2.06** | **1.01 - 4.30** | **0.050** |
|  | Seven months | 17 | 9 | 52.9 | 1.66 | 0.57 - 5.05 | 0.360 | 2.67 | 0.74 - 10.2 | 0.139 |

\*reference category
P-value calculated using the chi-square test
\*\*Excludes two unknowns
^Participants were asked which type of patient contact describes MOST of their current work (excludes five unknowns)

To broaden the analysis, multivariable logistic regression was repeated including participants that had grayzone results. The results of this analysis were similar to the results of the initial analysis and are presented in Appendix 4. Separate analysis was carried out to explore the association between participant characteristics and Abbott grayzone results; there was no significant association observed (data not presented).

## Discussion

### Seropositivity in relation to assay type: Abbott versus Roche – all participants

In agreement with recently published data (12), a considerably higher proportion of participants (more than double) had detectable antibodies on the Roche assay compared to the Abbott assay.

In terms of assay performance, according to the manufacturer’s instructions both assays perform with high sensitivity and high specificity. Applying the lower bounds of the confidence interval for specificity of the Abbott assay, a maximum false negative rate of 4.2% (n=15) could be expected, therefore 109 out of 124 negative test results among those previously diagnosed with COVID-19 (positive by PCR) cannot be explained by expected test performance; it is possible that SARS-CoV-2 IgG was absent among these individuals. Applying the lower bounds of the of the confidence interval for specificity of the Roche assay, a maximum false negative rate of 0.3% (n=1) could be expected, therefore 18 out of 19 negative test results among those previously diagnosed with COVID-19 (SARS-CoV-2 positive by PCR) cannot be explained by expected test performance. It is possible that some of these individuals did not mount a SARS-CoV-2 antibody response to their infection, however in the published literature the majority of patients have been shown to develop antibodies following natural infection (5) (21) (22), therefore it is likely that the main reason for lack of antibodies in this subset of participants was due to waning of antibody response that was not picked up on testing, particularly on the Abbott assay. Studies have shown that anti-nucleocapsid IgG antibodies may be less likely to develop than IgM antibodies; Zhao et al showed that at day 15 post infection 100.0% had detected total Ab, 94.3% had IgM compared to only 79.8% having IgG Ab (5). Peterson et al and Kaufman et al showed that IgG antibodies were not detected for between 6.3% and 9% of people in the first 2 weeks after PCR confirmation of infection (23) (24) and that this proportion rose to 14% by 99-121 days (24).

Lumley at al demonstrated stable anti-spike IgG antibodies but waning anti-nucleocapsid IgG antibodies over time among HCWs in the United Kingdom healthcare service (13), as did Pelleau et al. (25). This may in part explain the Abbott IgG assay’s relative performance in detecting past infection, compared to the Roche total antibody assay; the total antibody approach used in the Roche Elecsys system may result in improved and sustained sensitivity suitable for population seroprevalence studies.

Twenty-four participants had detectable antibodies on the Abbott assay but not on the Roche assay, of those three had detectable antibodies on the Wantai assay (one of whom had recent PCR-confirmed infection) and were presumably false negatives on Roche, possibly reflecting differences in test method and target between the Wantai and Roche assays. Among the remaining 21 that did not have detectable antibodies on either the Roche or the Wantai assay there were no participants that had previous PCR-confirmed SARS-CoV-2 infection. While it is possible that these were false positive results, interpretation of positive Abbott results for these 21 individuals is difficult as clinical significance and the duration of persistence of each type of SARS-CoV-2 antibody has yet to be fully elucidated.

### Does the Abbott grayzone add anything?

In October 2020, Abbott updated their guidance on their Architect SARS-CoV-2 IgG assay (Abbott Diagnostics Product Information Letter PI1060-2020) to include an optional editable “grayzone” with a S/C index range of 0.5 – 1.39 which they advise “must be interpreted by the clinician in the context of relevant clinical and laboratory information on the patient” (16). The grayzone accounts for a large proportion of the disagreement between the assays. The majority of grayzones were positive by Roche (67%) and Wantai (70%). However, interpretation of this suggested Abbott grayzone result in a clinical setting would not be straightforward and confirmatory testing would still be needed.

There was no obvious correlation between increasing or decreasing Abbott grayzone S/C index and the interpreted results from other assays. In an epidemiological study setting such as this, inclusion of all grayzone results as presumptive positives would lead to significant overestimation of seroprevalence. Studies such as ours could be used to estimate the proportion of grayzone results likely to be positive on other testing platforms, however the use of alternative assays with prolonged sensitivity over time would be superior if the primary purpose is to estimate infection ever. It is notable that while studies have shown good protection against reinfection over a six month period following PCR-confirmed COVID-19 infection (4), to the best of our knowledge no studies have yet compared antibody assays in terms of functional immunity.

### Seropositivity in relation to assay type – Roche versus Wantai assay

We found almost perfect agreement between the Roche and Wantai assays. A study comparing eight assays found these two assays to provide the highest sensitivities at 98 and 95 percent respectively (25). However, in our study the assay concordance should be interpreted with caution as selection of participants for additional testing on Wantai ELISA assay was heavily biased; only participants who had a positive or grayzone Abbott SARS-CoV-2 IgG result were selected for testing by Wantai assay. The small degree of discordance between these two assays could be due to a number of reasons. Although these two assays both measure SARS-CoV-2 total antibodies, they are based on different methodology (Roche Elecsys Anti-SARS-CoV2 Total AB is based on chemiluminescence and targets the nucleocapsid, whereas the Fortress Wantai Total AB assay is based on ELISA and targets the spike protein). Furthermore, there is a difference in expected performance of these assays in terms of specificity and sensitivity, according to manufacturer guidelines (19).

### Seropositivity over time (previously PCR-positive participants)

Of the subset of participants who had previous PCR-confirmed COVID-19 infection, the Roche assay performed better in terms of identifying these participants; 95% tested positive. Only half of these past infections were accurately identified by the Abbott assay. Harley et al. showed a much higher level of agreement between these two assays, though on a smaller number of samples than were tested in our study and at an earlier time point post PCRI-confirmation of infection (26). All previously PCR-positive participants in our study had detectable antibodies on the Wantai ELISA assay, but this subpopulation examination and its inherent selection bias limits strong conclusions from this data.

The majority of participants had a six-month interval between confirmed infection and antibody testing in our study, which correlated with the time period between the peak of the first wave of the pandemic in Ireland and the antibody testing in our study. Although pick-up of these was slightly lower on both assays than the overall pickup of previous infections occurring at any time, the Roche still performed significantly better than Abbott using the 6-month timeframe, identifying 93% versus 46% of infections that occurred six months prior.

The decay in seropositivity on Abbott in our cohort started at 21 weeks (150 days) after confirmed infection, but the small numbers of infection per week prior to this should be noted. This did not change with removal of participants who had no symptoms at the time of infection (Table 7). A study comparing four different serological assays, including the Abbott and Roche assays, showed a decline in the performance of the Abbott assay after 60 days, whereas antibodies were still detected on the Roche assay after 80 days (12). In contrast, another study of the Abbott assay showed a mean of 137 days to loss of positive antibody (13). Kumar et al. showed a much shorter duration of antibody detection using the Roche assay after confirmed SARS-CoV-2 infection in a small cohort of Indian HCWs; seropositivity halved by day 30-42 and fell to zero after 50 days (27). In a large US Study, Kaufman et al. showed a decline in IgG antibodies (including on Abbott) to 74% by 2 months after confirmed infection (24). Due to the small numbers of infection per week prior to the 21 –week time point in our study, it is possible that this drop-off in Abbott IgG positivity started at an earlier timepoint that was not picked up in our study. While we demonstrate good performance of Roche anti-nucleocapsid assay at extended time points, the comparison with other antigenic targets at time points even more distant from the infection remains obscure.

In a multivariable analysis, the risk of a negative result on the Abbott platform despite previous PCR-confirmed COVID-19 infection increased with interval between infection and antibody testing. Those who had moderate or severe symptoms were more likely to have retained IgG than those who only had mild symptoms. This was consistent with the findings of other researchers who found that IgG was better sustained in persons reporting significant symptoms compared to those who had mild or no symptoms (27) (28) (29). Participants of male sex and Asian ethnicity were also more likely to have sustained Abbott assay detected IgG. Kaufman et al. showed age and male sex to be associated with the probability of persistent IgG serology (24), as did a large Cochrane review conducted in April 2020 (31). Furthermore, Lumley at al. demonstrated that increasing age, Asian ethnicity and prior self-reported symptoms were independently associated with higher maximum anti-nucleocapsid IgG levels (13). This is in keeping with our findings on sex, ethnicity and prior self-reported symptoms, however we did not find any statistically significant correlation with age. The reason for this sustained IgG response in those of male sex is not yet clear but may be related to higher viral load, other indices of severity, or other unknown biological factors not included in this study.

Participants who had a six-month PCR to serology testing interval were twice as likely to be IgG seronegative when compared to participants who had a five-month testing interval. Those who had a seven-month testing were also more likely to be IgG seronegative when compared to those who had a five-month testing interval, but results were not significant. Our findings are consistent with the findings of other researchers who have demonstrated decline in IgG seropositivity over time, using the Abbott and other IgG antibody assays (7) (26). Further studies may provide better understanding of seropositivity and seroreversion over time, relating to specific assays or type of immunoglobulin measured.

We did not have enough participants with very recent infection to assess the ability of each assay to pick up early infection, however of the eight infections that occurred within four weeks of antibody testing, 8/9 were correctly identified by the Abbott assay and 6/9 were identified by the Roche assay. Higher numbers would be needed to compare these assays specifically in the early stages of infection.

Almost 5% of infections were not identified by either platform. The majority were distant infections (≥6 months ago), and therefore waning immunity may explain the seronegativity. One was a recent infection (PCR positive 24 days prior to serological testing) in a participant who had severe symptoms; this infection may have been too recent to have detectable antibodies on either assay, however four other participants with only minor symptoms and even more recent infections (PCR positive 12-17 days prior to serology), had antibodies detected on both Abbott and Roche assays. Overall, less of these participants with previous confirmed infection and negative serology on both platforms were symptomatic at the time of testing positive by PCR; other studies have shown those without symptoms to be both less likely to develop antibodies, and less likely to have persistent antibodies eight weeks post-infection (28). There were no other participant characteristics that were significantly associated with negative serology, nor were these participants quantitative results (Abbott S/C, Roche COI index) close to the positive result threshold for either assay. Both assays performed below their expected sensitivity based on manufacturers’ guidelines, likely due to the time-interval between infection and antibody testing. This should be considered in serological studies, as well as when using antibody testing in the setting of clinical care. As the pandemic progresses, further studies may highlight the sensitivity of different assays with more accuracy in relation to timing of testing.

## Limitations

Our study has several limitations. Firstly, the study was not designed with the primary objective of comparing the serological assays; two anti-nucleocapsid assays +/-additional testing with an anti-spike assay were used to maximise sensitivity in detection of SARS-CoV-2 antibodies. Secondly, information on COVID-19 symptoms and PCR test results were self-reported and thus could be biased. The dates of PCR-confirmed infection (also self-reported) may be inaccurate, furthermore the PCR cycle threshold (Ct) value which would have been a valuable addition to this study was not available. Other variables which would have been valuable to this study but were not available include participant co-morbidities. A small sample size for the 0-4-month PCR to serology test interval prevented meaningful analysis of seropositivity and seronegativity at the early stages post infection.

Our study focused on assays that have SARS CoV-2 nucleocapsid as an antigenic target with only a subpopulation assessed using a spike antibody assay. Given emerging evidence of differences in antibody decay related to the antigenic target a more complete assessment would have been desirable. Such approaches including parallel spike and nucleocapsid assessments may become more relevant in the era of SARS CoV-2 vaccination. In addition, we did not address the neutralising capacity of the antibodies measured, therefore conclusions about the functional consequences of the presence of such antibodies are limited.

## Conclusion and Recommendations

The Roche assay performed significantly better at picking up those who had ever had a confirmed COVID-19 infection, though both the Roche and Abbott assays were less sensitive than the manufacturers’ stated guidelines. Our study findings suggest that, of these two assays directed at anti-nucleocapsid antibodies, Roche is better suited to future population-based serological studies, due to maintained detection of total antibodies up to at least seven months after natural infection with SARS-CoV-2. While maximum sensitivity is achieved using multiple testing platforms, this is not always feasible or cost-effective, and the overall seroprevalence results of our study would have been unchanged if only the Roche assay was used. While we demonstrate good performance of Roche anti-nucleocapsid assay at extended time points, the comparison with other antigenic targets at time points even more distant from the infection remains obscure. The anti-spike assay (Wantai) performed very well on the subset of samples analysed, but further studies are needed to show if it maintains its sensitivity compared to Roche on an unbiased selection of samples.

The risk of a negative antibody result on the Abbott assay despite previous PCR-confirmed SARS-C0V-2 infection increased with increasing interval between infection and time of serological testing; the drop-off was noted 21 weeks/150 days after confirmed SARS-CoV-2 infection. Those of male sex, Asian ethnicity and those with moderate to severe symptoms were more likely to retain IgG on the Abbott assay. Samples with a S/C index within the Abbott grayzone still require additional testing on an alternative assay, and the numerical value within the grayzone S/C index range does not predict the result of confirmatory testing. From our limited numbers of recent infections, it may be possible that the Abbott assay picked up early infection quicker than the Roche assay, but our numbers were too small to determine this. If further studies were to confirm this finding, then the Abbott assay may have a clinical role in early infection where serial PCR fails to diagnose SARS-CoV-2 infection, but clinical suspicion remains high.

Our study adds to the growing literature on serological assays for population-based studies. Further data comparing antibody assays for the detection of antibodies to SARS-CoV-2 are needed to guide the most appropriate use of certain assays in any given situation, especially where use of dual or multiple assays is not feasible or affordable. With the recent introduction of widespread vaccination it is yet to be determined if measuring antibody response to vaccination is meaningful and cost-effective, and if so which assays are superior. Further studies are also needed to delineate the relationship between serological response and functional immunity to SARS-CoV-2 infection, both following vaccination and natural infection. Assays with prolonged sensitivity are likely to be more valuable as the pandemic continues.

## Data Availability

Dataset has not been made available as may identify individual participants. Dataset can be made available upon reasonable request to the Principal Investigator.

## Acknowledgements

We would like to acknowledge the study steering group who planned the study and critically evaluated this manuscript, the study team who coordinated the running of the study in each hospital, the hospital management at both sites for their support for the study, and the staff of both hospitals who participated. We would especially like to acknowledge the phlebotomy departments in each hospital for facilitating the sampling of almost 6000 participants, the microbiology, virology and biochemistry laboratories in each hospital for processing the samples on two different assays, the National Virus Reference Laboratory of Ireland for additional testing and the human resources department for their help with denominator data.

## Ethical Approval

Ethical approval was obtained from the National Research Ethics Committee for COVID-19 in Ireland (20-NREC-COV-101).

## Appendices

### Appendix 1A Characteristics of participants tested, by serology assay

|  |  | Abbott SARS-CoV-2 IgG |  | Roche Anti-SARS-CoV-2 |  | Wantai SARS-CoV-2 Antibody ELISA |  | Total participants |  |
| --- | --- | --- | --- | --- | --- | --- | --- | --- | --- |
| Total participants tested |  | (n=5,788) |  | (n=5,787) |  | (n=485) |  | (n=5,788) |  |
|  |  | n | % | n | % | n | % | n | % |
| **Age group (years)** | 18-29 | 1350 | 23.3 | 1350 | 23.3 | 139 | 28.7 | 1350 | 23.3 |
|  | 30-39 | 1617 | 27.9 | 1617 | 27.9 | 139 | 28.7 | 1617 | 27.9 |
|  | 40-49 | 1516 | 26.2 | 1515 | 26.2 | 111 | 22.9 | 1516 | 26.2 |
|  | 50-59 | 1001 | 17.3 | 1001 | 17.3 | 68 | 14.0 | 1001 | 17.3 |
|  | 60+ | 304 | 5.3 | 304 | 5.3 | 28 | 5.8 | 304 | 5.3 |
| **Sex** | Female | 4478 | 77.4 | 4478 | 77.4 | 349 | 72.0 | 4478 | 77.4 |
|  | Male | 1309 | 22.6 | 1308 | 22.6 | 136 | 28.0 | 1309 | 22.6 |
|  | Unknown | 1 | 0.0 | 1 | 0.0 | 0 | 0.0 | 1 | 0.0 |
| **Ethnicity** | Irish | 4444 | 76.8 | 4444 | 76.8 | 313 | 64.5 | 4444 | 76.8 |
|  | Any other white background | 552 | 9.5 | 551 | 9.5 | 54 | 11.1 | 552 | 9.5 |
|  | Any Asian background | 577 | 10.0 | 577 | 10.0 | 101 | 20.8 | 577 | 10.0 |
|  | Any African or black background | 113 | 2.0 | 113 | 2.0 | 13 | 2.7 | 113 | 2.0 |
|  | Other | 101 | 1.7 | 101 | 1.7 | 4 | 0.8 | 101 | 1.7 |
|  | Unknown | 1 | 0.0 | 1 | 0.0 | 0 | 0 | 1 | 0.0 |
| **Role** | Nursing/midwifery | 2064 | 35.7 | 2064 | 35.7 | 230 | 47.4 | 2064 | 35.7 |
|  | Allied health | 1091 | 18.8 | 1091 | 18.9 | 57 | 11.8 | 1091 | 18.8 |
|  | Medical/dental | 983 | 17.0 | 982 | 17.0 | 93 | 19.2 | 983 | 17.0 |
|  | Admin | 803 | 13.9 | 803 | 13.9 | 33 | 6.8 | 803 | 13.9 |
|  | General support | 434 | 7.5 | 434 | 7.5 | 21 | 4.3 | 434 | 7.5 |
|  | Health care assistant | 286 | 4.9 | 286 | 4.9 | 45 | 9.3 | 286 | 4.9 |
|  | Other | 127 | 2.2 | 127 | 2.2 | 6 | 1.2 | 127 | 2.2 |

### Appendix 1B COVID-19 related characteristics of participants tested, by serology assay

|  |  | Abbott SARS-CoV-2<br>IgG |  | Roche Anti-SARS-CoV-2 |  | Wantai SARS-CoV-2<br>Antibody ELISA |  | Total participants |  |
| --- | --- | --- | --- | --- | --- | --- | --- | --- | --- |
| Total participants tested |  | (n = 5,788) |  | (n = 5,787) |  | (n = 485) |  | (n = 5,788) |  |
|  |  | n | % | n | % | n | % | n | % |
| **Close contact with a case of COVID-19** | Yes | 1705 | 29.5 | 1704 | 29.4 | 258 | 53.2 | 1705 | 29.5 |
|  | No | 4071 | 70.3 | 4071 | 70.3 | 225 | 46.4 | 4071 | 70.3 |
|  | Unknown | 12 | 0.2 | 12 | 0.2 | 2 | 0.4 | 12 | 0.2 |
| **Main type of patient contact\*** | Daily contact with known/suspected COVID-19 | 903 | 15.6 | 902 | 15.6 | 109 | 22.5 | 903 | 15.6 |
|  | Daily contact with patients without COVID-19 | 3245 | 56.1 | 3245 | 56.1 | 299 | 61.6 | 3245 | 56.1 |
|  | No patient contact | 1635 | 28.2 | 1635 | 28.3 | 77 | 15.9 | 1635 | 28.2 |
|  | Unknown | 5 | 0.1 | 5 | 0.1 | 0 | 0.0 | 5 | 0.1 |
| **Previous COVID-19 symptoms** | No symptoms | 2869 | 49.6 | 2868 | 49.6 | 102 | 21.0 | 2869 | 49.6 |
|  | Had symptoms | 2911 | 50.3 | 2911 | 50.3 | 381 | 78.6 | 2911 | 50.3 |
|  | Unknown | 8 | 0.1 | 8 | 0.1 | 2 | 0.4 | 8 | 0.1 |
| **Severity of symptoms** | Minor symptoms | 2159 | 74.2 | 2159 | 74.2 | 193 | 50.7 | 2159 | 74.2 |
|  | Significant symptoms | 701 | 24.1 | 701 | 24.1 | 161 | 42.3 | 701 | 24.1 |
|  | Severe symptoms (hospitalised) | 51 | 1.8 | 51 | 1.8 | 27 | 7.1 | 51 | 1.8 |
| **Previous COVID-19 PCR test** | Yes | 2779 | 48.0 | 2778 | 48.0 | 380 | 78.4 | 2779 | 48.0 |
|  | No | 3003 | 51.9 | 3003 | 51.9 | 105 | 21.6 | 3003 | 51.9 |
|  | Unknown | 6 | 0.1 | 6 | 0.1 | 0 | 0.0 | 6 | 0.1 |
| **Previous positive COVID-19** | Yes | 367 | 6.3 | 367 | 6.3 | 259 | 53.4 | 367 | 6.3 |
|  | No | 5415 | 93.6 | 5414 | 93.6 | 226 | 46.6 | 5415 | 93.6 |
|  | Unknown | 6 | 0.1 | 6 | 0.1 | 0 | 0.0 | 6 | 0.1 |

\*Participants were asked which one describes MOST of their current work

### Appendix 2 Characteristics of participants who had a previously PCR-confirmed infection and were negative by serology testing (n=17)

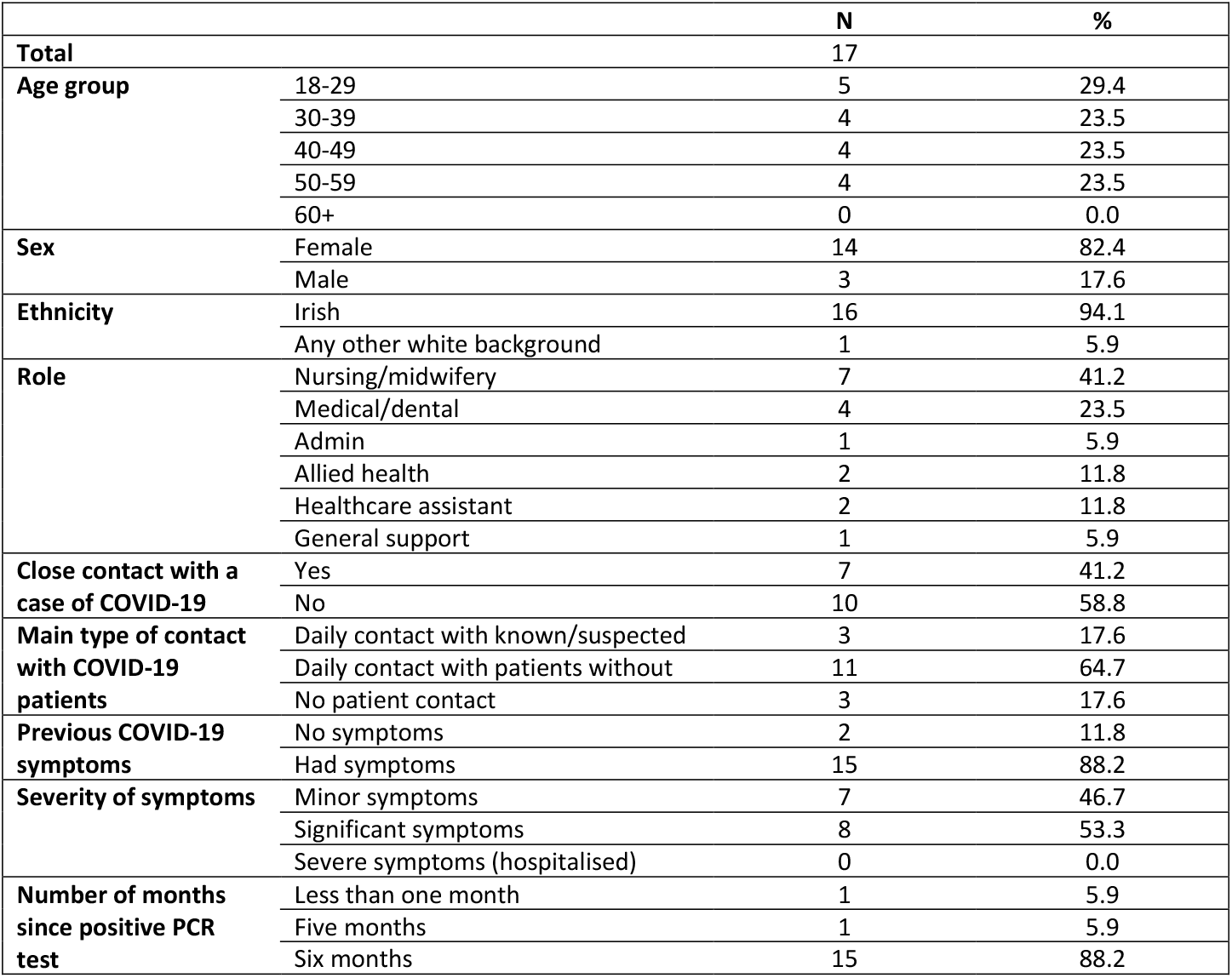

|  |  | **N** | **%** |
| --- | --- | --- | --- |
| **Total** |  | 17 |  |
| **Age group** | 18-29 | 5 | 29.4 |
|  | 30-39 | 4 | 23.5 |
|  | 40-49 | 4 | 23.5 |
|  | 50-59 | 4 | 23.5 |
|  | 60+ | 0 | 0.0 |
| **Sex** | Female | 14 | 82.4 |
|  | Male | 3 | 17.6 |
| **Ethnicity** | Irish | 16 | 94.1 |
|  | Any other white background | 1 | 5.9 |
| **Role** | Nursing/midwifery | 7 | 41.2 |
|  | Medical/dental | 4 | 23.5 |
|  | Admin | 1 | 5.9 |
|  | Allied health | 2 | 11.8 |
|  | Healthcare assistant | 2 | 11.8 |
|  | General support | 1 | 5.9 |
| **Close contact with a case of COVID-19** | Yes | 7 | 41.2 |
|  | No | 10 | 58.8 |
| **Main type of contact with COVID-19 patients** | Daily contact with known/suspected | 3 | 17.6 |
|  | Daily contact with patients without | 11 | 64.7 |
|  | No patient contact | 3 | 17.6 |
| **Previous COVID-19 symptoms** | No symptoms | 2 | 11.8 |
|  | Had symptoms | 15 | 88.2 |
| **Severity of symptoms** | Minor symptoms | 7 | 46.7 |
|  | Significant symptoms | 8 | 53.3 |
|  | Severe symptoms (hospitalised) | 0 | 0.0 |
| **Number of months since positive PCR test** | Less than one month | 1 | 5.9 |
|  | Five months | 1 | 5.9 |
|  | Six months | 15 | 88.2 |

### Appendix 3A Characteristics of participants who had a previously PCR-confirmed infection and their test results on Abbott SARS-CoV-2 IgG (n=367)

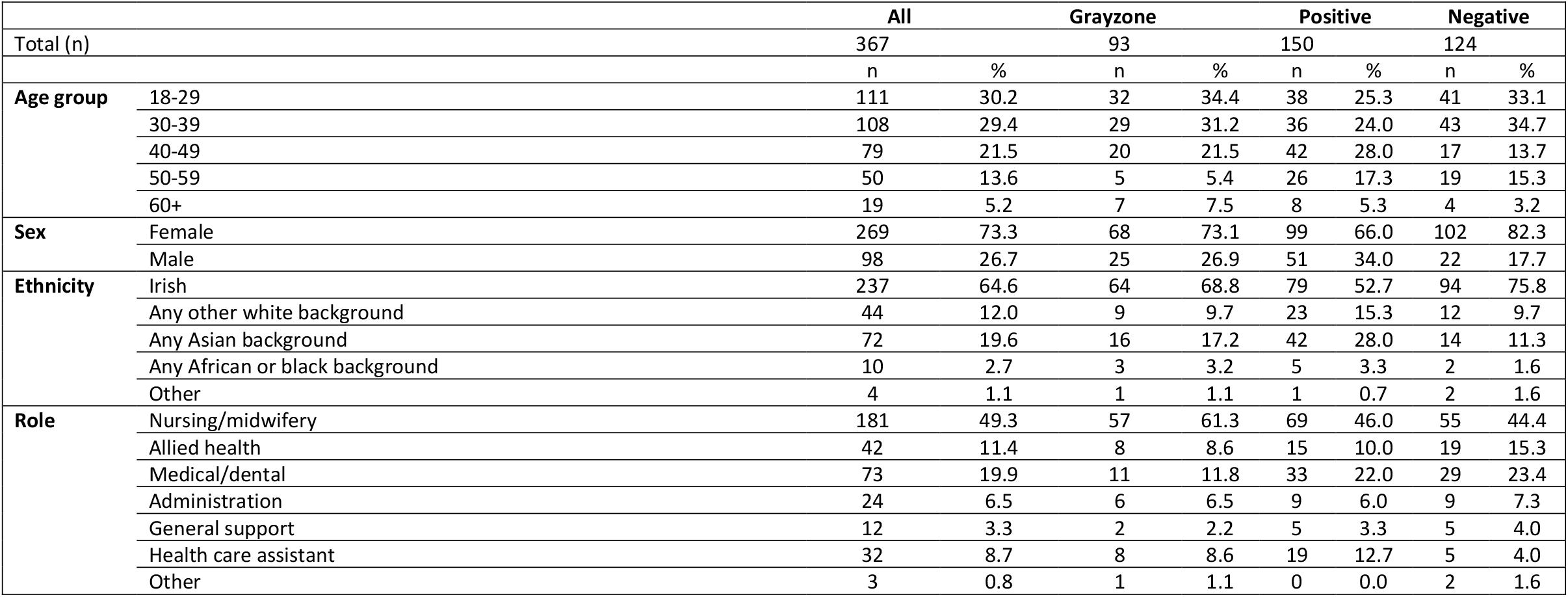

### Appendix 3B COVID-19 related characteristics of participants who had a previously PCR-confirmed infection and their test results on Abbott SARS-CoV-2 IgG (n=367)

|  |  | All |  | Grayzone |  | Positive |  | Negative |  |
| --- | --- | --- | --- | --- | --- | --- | --- | --- | --- |
| Total (n) |  | 367 |  | 93 |  | 150 |  | 124 |  |
|  |  | n | % | n | % | n | % | n | % |
| **Close contact with a case of COVID-19** | Yes | 211 | 57.5 | 56 | 60.2 | 90 | 60.0 | 65 | 52.4 |
|  | No | 154 | 42.0 | 37 | 39.8 | 58 | 38.7 | 59 | 47.6 |
|  | Unknown | 2 | 0.5 | 0 | 0.0 | 2 | 1.3 | 0 | 0.0 |
| **Main type of patient contact\*** | Daily contact with known/suspected COVID-19 patients | 88 | 24.0 | 22 | 23.7 | 40 | 26.7 | 26 | 21.0 |
|  | Daily contact with patients without COVID-19 | 218 | 59.4 | 56 | 60.2 | 88 | 58.7 | 74 | 59.7 |
|  | No patient contact | 61 | 16.6 | 15 | 16.1 | 22 | 14.7 | 24 | 19.4 |
| **Symptoms at time of previous PCR test** | No symptoms | 7 | 1.9 | 1 | 1.1 | 3 | 2.0 | 3 | 2.4 |
|  | Had symptoms | 358 | 97.5 | 92 | 98.9 | 145 | 96.7 | 121 | 97.6 |
|  | Unknown | 2 | 0.5 | 0 | 0.0 | 2 | 1.3 | 0 | 0.0 |
| **Severity of symptoms** | Minor symptoms | 151 | 41.1 | 38 | 43.2 | 51 | 35.2 | 62 | 51.2 |
|  | Significant symptoms | 178 | 48.5 | 48 | 54.5 | 75 | 51.7 | 55 | 45.5 |
|  | Severe symptoms (hospitalised) | 29 | 7.9 | 6 | 6.8 | 19 | 13.1 | 4 | 3.3 |
| **Number of months since positive PCR test** | Less than one month | 8 | 2.2 | 1 | 1.1 | 6 | 4.0 | 1 | 0.8 |
|  | One month | 8 | 2.2 | 0 | 0.0 | 8 | 5.3 | 0 | 0.0 |
|  | Two months | 4 | 1.1 | 0 | 0.0 | 4 | 2.7 | 0 | 0.0 |
|  | Three months | 1 | 0.3 | 0 | 0.0 | 1 | 0.7 | 0 | 0.0 |
|  | Four months | 12 | 3.3 | 2 | 2.2 | 10 | 6.7 | 0 | 0.0 |
|  | Five months | 85 | 23.2 | 28 | 30.1 | 34 | 22.7 | 23 | 18.5 |
|  | Six months | 222 | 60.5 | 54 | 58.1 | 77 | 51.3 | 91 | 73.4 |
|  | Seven months | 25 | 6.8 | 8 | 8.6 | 8 | 5.3 | 9 | 7.3 |
|  | Unknown | 2 | 0.5 | 0 | 0.0 | 2 | 1.3 | 0 | 0.0 |
| **Number of months since onset of symptoms\*\*** | Less than one month | 5 | 1.5 | 1 | 1.1 | 3 | 2.2 | 1 | 0.9 |
|  | One month | 8 | 2.4 | 0 | 0.0 | 8 | 5.8 | 0 | 0.0 |
|  | Two months | 3 | 0.9 | 0 | 0.0 | 3 | 2.2 | 0 | 0.0 |
|  | Three months | 1 | 0.3 | 0 | 0.0 | 1 | 0.7 | 0 | 0.0 |
|  | Four months | 12 | 3.5 | 2 | 2.3 | 10 | 7.2 | 0 | 0.0 |
|  | Five months | 78 | 22.9 | 25 | 28.4 | 32 | 23.2 | 21 | 18.4 |
|  | Six months | 210 | 61.8 | 52 | 59.1 | 73 | 52.9 | 85 | 74.6 |
|  | Seven months | 23 | 6.8 | 8 | 9.1 | 8 | 5.8 | 7 | 6.1 |

\*Participants were asked which one describes MOST of their current work \*\* excludes 27 participants who were not symptomatic at the time of their positive PCR test

### Appendix 4 Factors associated with Abbott SARS-CoV-2 IgG seronegativity, among participants who had a previously PCR-confirmed infection, including those who had a grayzone result (n=367)

|  |  | All<br>(n) | Negative<br>(n) | Negative<br>% | Unadjusted<br>OR | 95% CI | P-value | Adjusted<br>OR | 95% CI | P-value |
| --- | --- | --- | --- | --- | --- | --- | --- | --- | --- | --- |
| **Total** |  | 367 | 124 | 33.8 |  |  |  |  |  |  |
| **Age group** | 18-29 | 111 | 41 | 36.9 | * |  |  | * |  |  |
|  | 30-39 | 108 | 43 | 39.8 | 1.13 | 0.66 – 1.95 | 0.66 | 0.96 | 0.51 - 1.8 | 0.893 |
|  | 40-49 | 79 | 17 | 21.5 | 0.47 | 0.24 – 0.89 | 0.02 | 0.44 | 0.21 - 0.92 | 0.031 |
|  | 50-59 | 50 | 19 | 38.0 | 1.05 | 0.52 – 2.08 | 0.89 | 1.05 | 0.47 - 2.36 | 0.897 |
|  | 60+ | 19 | 4 | 21.1 | 0.46 | 0.12 – 1.35 | 0.18 | 0.32 | 0.08 - 1.11 | 0.090 |
| **Sex** | Female | 269 | 102 | 37.9 | * |  |  | * |  |  |
|  | Male | 98 | 22 | 22.4 | 0.47 | 0.28 – 0.81 | 0.00 | 0.38 | 0.21 - 0.68 | 0.001 |
| **Ethnicity** | Irish | 237 | 94 | 39.7 | * |  |  | * |  |  |
|  | Any other white background | 44 | 12 | 27.3 | 0.57 | 0.27 – 1.14 | 0.12 | 0.64 | 0.29 - 1.39 | 0.273 |
|  | Any Asian background | 72 | 14 | 19.4 | 0.37 | 0.19 – 0.68 | 0.00 | 0.41 | 0.19 - 0.81 | 0.012 |
|  | Any African/black background | 10 | 2 | 20.0 | 0.38 | 0.05 – 1.56 | 0.22 | 0.48 | 0.07 - 2.29 | 0.392 |
|  | Other | 4 | 2 | 50.0 | 1.52 | 0.18 – 12.9 | 0.67 | 1.53 | 0.15 - 15.7 | 0.705 |
| **Close contact**<br>**with a case of** | Yes | 211 | 65 | 30.8 | 0.72 | 0.46 – 1.11 | 0.14 | 0.67 | 0.4 - 1.12 | 0.127 |
|  | No | 154 | 59 | 38.3 | * |  |  | * |  |  |
| **Main type of**<br>**patient contact^** | No patient contact | 61 | 24 | 39.3 | * |  |  | * |  |  |
|  | Daily contact non-COVID-19 | 218 | 74 | 33.9 | 0.79 | 0.44 – 1.43 | 0.43 | 0.78 | 0.4 - 1.56 | 0.484 |
|  | Daily contact known or suspected | 88 | 26 | 29.5 | 0.65 | 0.21 – 0.32 | 0.21 | 0.84 | 0.38 - 1.89 | 0.679 |
| **Severity of**<br>**symptoms\*\*** | No symptoms | 7 | 3 | 42.9 | 1.08 | 0.21 – 5.05 | 0.92 | 1.07 | 0.17 - 6.16 | 0.942 |
|  | Minor symptoms | 151 | 62 | 41.1 | * |  |  | * |  |  |
|  | Significant symptoms | 178 | 55 | 30.9 | 0.64 | 0.41 – 1.01 | 0.05 | 0.58 | 0.34 - 0.96 | 0.037 |
|  | Severe symptoms (hospitalised) | 29 | 4 | 13.8 | 0.23 | 0.06 – 0.63 | 0.00 | 0.26 | 0.07 - 0.76 | 0.023 |
| **HCW Role** | Administration | 24 | 9 | 37.5 | * |  |  |  |  | not entered |
|  | Allied health care | 42 | 19 | 45.2 | 1.38 | 0.49 – 3.94 | 0.54 |  |  |  |
|  | General Support | 12 | 5 | 41.7 | 1.19 | 0.28 – 4.92 | 0.80 |  |  |  |
|  | Healthcare assistant | 32 | 5 | 15.6 | 0.31 | 0.08 – 1.06 | 0.06 |  |  |  |
|  | Medical/Dental | 73 | 29 | 39.7 | 1.10 | 0.43 – 2.9 | 0.84 |  |  |  |
|  | Nursing/midwifery | 181 | 55 | 30.4 | 0.73 | 0.31 – 1.83 | 0.48 |  |  |  |
|  | Other | 3 | 2 | 66.7 | 3.33 | 0.28 – 78.0 | 0.35 |  |  |  |
| **Number of** | Less than one month | 8 | 1 | 12.5 | NA | NA | NA | 0.23 | 0.01 - 1.6 | 0.204 |
| **months since PCR positive test\*\*** | One month | 8 | 0 | 0.0 | NA | NA | NA | NA | NA | NA |
|  | Two months | 4 | 0 | 0.0 | NA | NA | NA | NA | NA | NA |
|  | Three months | 1 | 0 | 0.0 | NA | NA | NA | NA | NA | NA |
|  | Four months | 12 | 0 | 0.0 | NA | NA | NA | NA | NA | NA |
|  | Five months | 85 | 23 | 27.1 | * |  |  | * |  |  |
|  | Six months | 222 | 91 | 41.0 | 1.87 | 1.09 – 3.29 | 0.02 | 1.95 | 1.07 - 3.64 | 0.032 |
|  | Seven months | 25 | 9 | 36.0 | 1.52 | 0.57 – 3.87 | 0.38 | 2.07 | 0.72 - 5.9 | 0.172 |

\*reference category P-value calculated using the chi-square test \*\*Excludes two unknowns ^Participants were asked which type of patient contact describes MOST of their current work (excludes five unknowns)

